# Pantoprazole Use in Invasively Ventilated Patients With Septic Shock: A Protocol and Statistical Analysis Plan

**DOI:** 10.64898/2026.04.27.26351851

**Authors:** Akira Kuriyama, Diane Heels-Ansdell, Shannon M. Fernando, Neill K.J. Adhikari, Francois Lamontagne, Bijan Teja, Kimberly Anne Lewis, Bram Rochwerg, Karrilloi Laiya Carayannopoulos, Gloria Vazquez-Grande, Lauralyn McIntyre, Kimia Honarmand, Dipayan Chaudhuri, Mette Krag, Nicole Zytaruk, Deborah Cook, the Canadian Critical Care Trials Group

## Abstract

**Background:** Sepsis is a recognized risk factor for upper gastrointestinal bleeding, yet sepsis-specific randomized evidence informing stress ulcer prophylaxis remains limited.

**Objective:** To describe the rationale, methods, and statistical analysis plan for a post hoc subgroup analysis evaluating pantoprazole versus placebo in invasively ventilated critically ill adults with septic shock enrolled in the REVISE trial (NCT03374800).

**Methods:** This study will be a post hoc extended subgroup analysis of the international, blinded, randomized REVISE trial, which enrolled 4,821 mechanically ventilated adults in 68 ICUs across 8 countries. Patients were randomized to intravenous pantoprazole 40 mg once daily or placebo during invasive mechanical ventilation. Septic shock will be defined as receipt of vasopressors or inotropes at baseline together with an admitting diagnosis of infection according to APACHE III diagnostic categories.

**Results:** The primary efficacy outcome will be clinically important upper gastrointestinal bleeding in the ICU within 90 days after randomization, and the primary safety outcome will be all-cause mortality within 90 days. Additional trial outcomes will include patient-important upper gastrointestinal bleeding, ventilator-associated pneumonia, *Clostridioides difficile* infection during hospitalization, new renal replacement therapy, mortality in the ICU and hospital, and duration of ICU and hospital stay. Analyses will be adjusted for prehospital acid suppression; the mortality analyses will be additionally adjusted for APACHE II score.

**Conclusion:** This protocol and statistical analysis plan describes an evaluation of the efficacy and safety of pantoprazole in patients with septic shock within a large randomized trial dataset.

## Introduction

Sepsis is defined as life-threatening organ dysfunction caused by a dysregulated host response to infection (1). Sepsis may progress to septic shock, which is associated with higher risk of morbidity and mortality, with hospital mortality rates as high as 40% (2). Worldwide, sepsis and septic shock remain a leading cause of death, particularly in low- and middle-income countries (3, 4). One complication among patients with sepsis is upper gastrointestinal bleeding (5, 6). This may be due to a variety of factors, including inflammation, splanchnic hypoperfusion, and ischemia-reperfusion injury, which impairs the integrity of the protective gastric mucosal lining (7, 8). The development of gastrointestinal bleeding may be associated with worse clinical outcomes – longer length of ICU stay and higher mortality - in patients with sepsis (7) as well as patients without sepsis (9). Therefore, stress ulcer prophylaxis (SUP) is commonly prescribed to reduce the risk of upper gastrointestinal bleeding in patients with sepsis and septic shock; however, it is unclear whether SUP may confer lesser, similar, or greater benefit for patients specifically with septic shock compared to other conditions.

Several large observational studies have examined the effect of SUP in patients with sepsis using administrative or electronic health database, the results of which are conflicting in terms of impact on upper gastrointestinal bleeding, infectious complications, and mortality (10-14). In two recent international trials, the proton pump inhibitor (PPI) pantoprazole compared to no SUP reduced the incidence of clinically important upper gastrointestinal bleeding (15, 16), and reduced patient important bleeding (17) in one of these trials (15); however, these benefits have not translated into improved survival (18).

Clinical practice guideline recommendations for SUP have changed over time for patients with sepsis and septic shock. In the 2012 Surviving Sepsis Campaign Guidelines, acid suppression was recommended for patients with severe sepsis or septic shock who had risk factors for gastrointestinal bleeding (19), extrapolating from general ICU populations. The 2016 guidelines recommended SUP for patients with risk factors, driven by consistent evidence of reduced clinically important upper gastrointestinal bleeding (20). However, the moderate quality of the supporting evidence and the lack of sepsis-specific randomized trials was noted, marking a shift toward a more selective approach. The 2021 guidelines no longer supported routine SUP for all patients with sepsis or septic shock; a weak recommendation supported SUP only for patients at increased risk of upper gastrointestinal bleeding (21). This was also reflected in the risk-based approach emphasized in the 2024 SCCM/ASHP guidelines (22). The 2026 guidelines made a conditional recommendation, suggesting SUP with PPI over no SUP in adults with sepsis or septic shock, while maintaining an individualized, risk⍰based approach (23).

Given the absence of sepsis-specific randomized trial evidence on SUP in the context of guideline recommendations based on indirect observational data, this study will examine the effect of pantoprazole in the subgroup of critically ill patients with septic shock enrolled in a large international SUP trial.

## Methods

This will be a post hoc subgroup analysis of the Re-Evaluating the Inhibition of Stress Erosions (REVISE) trial (15). REVISE (ClinicalTrials.gov, NCT03374800) was a randomized, blinded, placebo-controlled trial of 4,821 invasively ventilated patients in 68 ICUs in 8 countries which evaluated the efficacy and safety of pantoprazole for SUP (24). Adults aged ≥18 years admitted to an ICU and expected to remain mechanically invasively ventilated beyond the day after enrollment were eligible. Exclusion criteria were invasive ventilation for >72 hours before screening, receipt of ≥1 day of acid-suppressive therapy in the ICU, dual antiplatelet therapy or combined antiplatelet and therapeutic anticoagulation, active upper gastrointestinal bleeding, and contraindications to acid-suppressive therapy. Patients were randomly assigned to receive intravenous pantoprazole 40 mg once daily or placebo during invasive mechanical ventilation in the ICU for up to 90 days or until the discontinuation of invasive ventilation, or death, whichever came first; study drug was discontinued if contraindications or clinical indications for open-label PPI arose. Patients were enrolled from July 9, 2019, to October 30, 2023. REVISE demonstrated that pantoprazole significantly reduced clinically important and patient important upper gastrointestinal bleeding without impacting mortality or other clinical outcomes (15).

To address the objectives of this study, we will focus on the subgroup of patients with septic shock at baseline, defined as Acute Physiology and Chronic Health Evaluation (APACHE) III diagnostic categories of infection, which included pneumonia (bacterial, viral [including COVID-19], or fungal), urinary sepsis, gastrointestinal perforation or obstruction, cholecystitis, skin and soft tissue infection, or sepsis of another origin (e.g., meningitis) combined with receipt of vasopressors or inotropes at baseline. This operational definition of septic shock was based on variables used in REVISE. The non-septic shock group comprised all other enrolled patients who did not meet the foregoing criteria.

The primary efficacy outcome will be clinically important upper gastrointestinal bleeding occurring in the ICU within 90 days after randomization, as defined in the primary trial. Specifically, this was overt upper gastrointestinal bleeding accompanied by hemodynamic compromise or resulting in therapeutic interventions in the ICU, including a decrease in hemoglobin of ≥2 g/dL within 24 hours, transfusion of ≥2 units of packed red blood cells, hypotension or initiation of vasopressors or inotropes, or performance of an invasive therapeutic procedure, or leading to ICU readmission during the index hospitalization. All suspected events were adjudicated in duplicate by calibrated blinded physicians. The primary safety outcome will be death from any cause within 90 days.

This extended subgroup analysis will include secondary trial outcomes (ventilator-associated pneumonia, *Clostridioides difficile* infection during hospitalization, patient-important upper gastrointestinal bleeding, new renal replacement therapy, and ICU and hospital mortality) and tertiary trial outcomes (duration of ICU and hospital stay). Patient-important upper gastrointestinal bleeding is defined based on a prior mixed-methods study involving ICU survivors and their families (17), and included bleeding events that required at least one blood transfusion, vasopressor therapy, diagnostic endoscopy, computed tomographic angiography, or surgery, or that resulted in death, disability, or prolonged hospitalization (25).

### Analysis

Of the original sample size of 4,821 patients, 1,072 (22.2%) had septic shock according to our definition and 3,749 (77.8%) did not. Patients will be evaluated in the randomized group to which they were assigned according to the intention-to-treat analysis. We will perform Cox proportional-hazards analyses for the primary efficacy and safety outcomes and for the evaluation of dichotomous secondary outcomes, with adjustment for receipt of prehospital acid suppression (**Table 1**). As per the main trial report (15), the mortality outcome at all time points (in-ICU, in-hospital, and 90-day) will be additionally adjusted for baseline illness severity using the APACHE II score (range, 0–71, with higher scores indicating greater risk of death). Results will be reported as hazard ratios (HRs) and 95% confidence intervals (CIs). Graphical approaches will be used to assess the proportional hazards assumption for Cox regression. For the skewed outcomes of ICU and hospital length of stay, we will perform linear regression with the log-transformed length of stay as the dependent variable. Denominators will represent the number of patients with full follow-up data for each outcome.

**Table 1.**
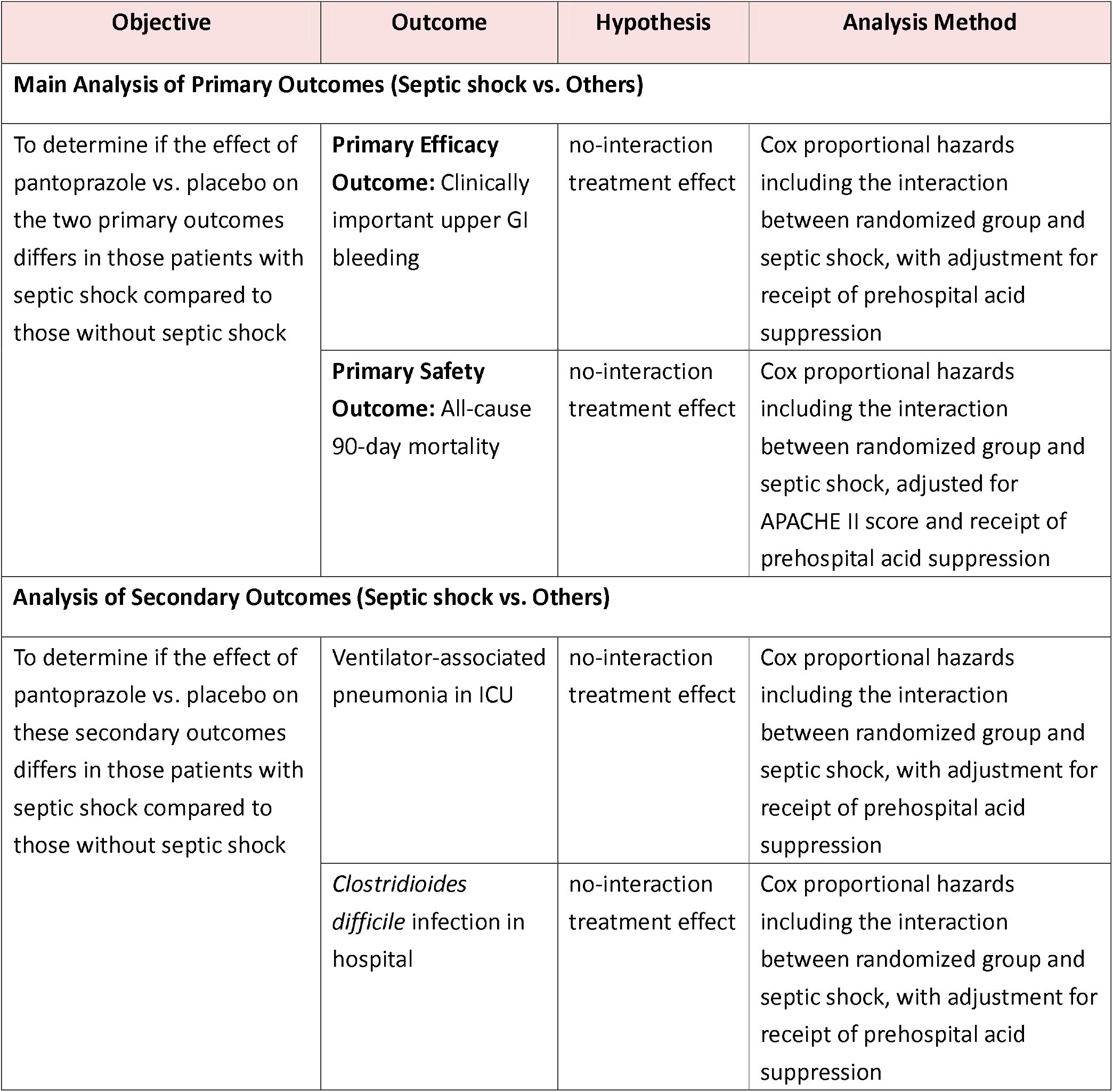
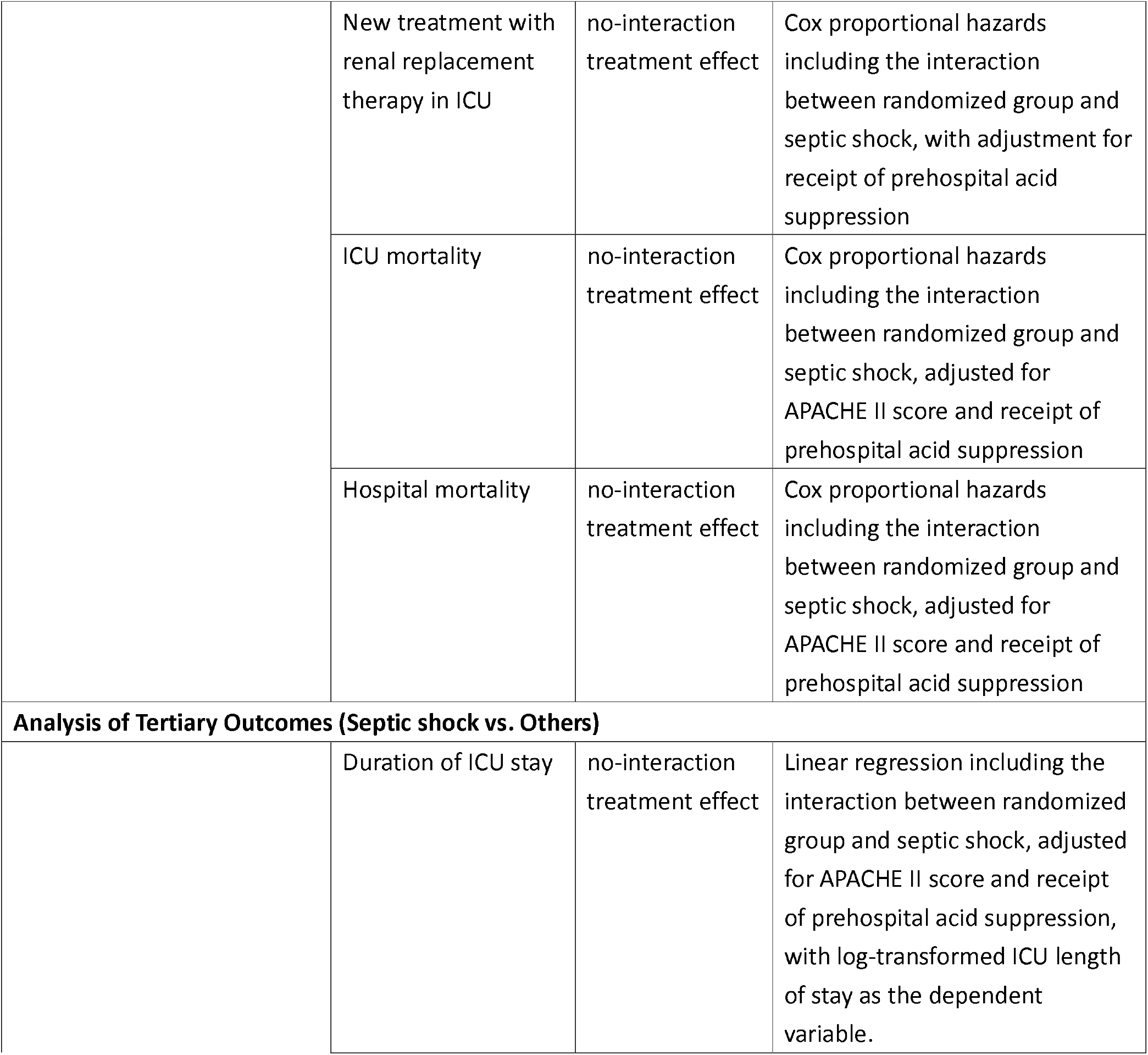
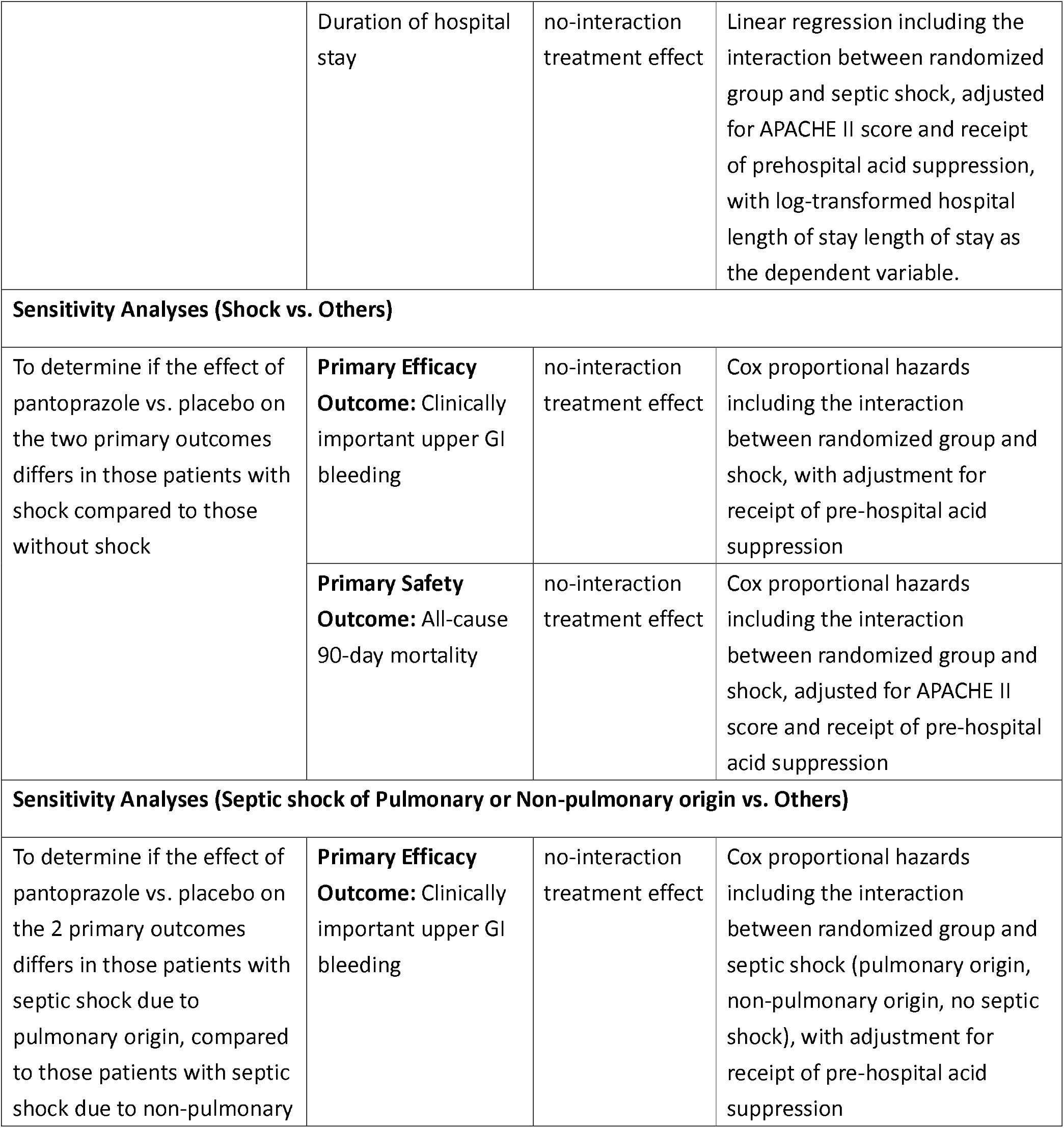
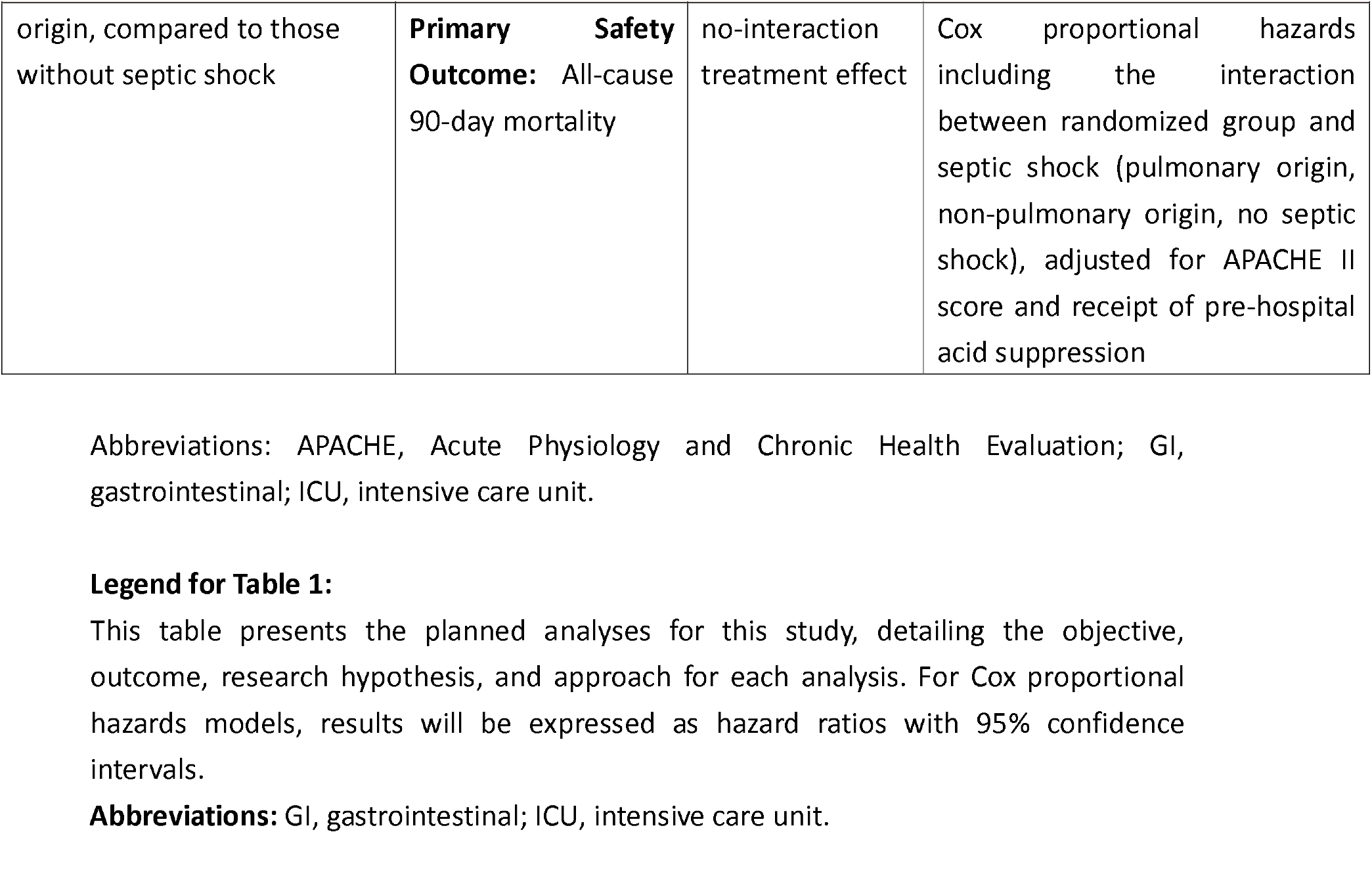
Summary of Objective, Hypothesis and Analysis Plan for Each Outcome.

| Objective | Outcome | Hypothesis | Analysis Method |
| --- | --- | --- | --- |
| **Main Analysis of Primary Outcomes (Septic shock vs. Others)** |  |  |  |
| To determine if the effect of pantoprazole vs. placebo on the two primary outcomes differs in those patients with septic shock compared to those without septic shock | **Primary Efficacy Outcome:** Clinically important upper GI bleeding | no-interaction treatment effect | Cox proportional hazards including the interaction between randomized group and septic shock, with adjustment for receipt of prehospital acid suppression |
|  | **Primary Safety Outcome:** All-cause 90-day mortality | no-interaction treatment effect | Cox proportional hazards including the interaction between randomized group and septic shock, adjusted for APACHE II score and receipt of prehospital acid suppression |
| **Analysis of Secondary Outcomes (Septic shock vs. Others)** |  |  |  |
| To determine if the effect of pantoprazole vs. placebo on these secondary outcomes differs in those patients with septic shock compared to those without septic shock | Ventilator-associated pneumonia in ICU | no-interaction treatment effect | Cox proportional hazards including the interaction between randomized group and septic shock, with adjustment for receipt of prehospital acid suppression |
|  | *Clostridioides difficile* infection in hospital | no-interaction treatment effect | Cox proportional hazards including the interaction between randomized group and septic shock, with adjustment for receipt of prehospital acid suppression |
|  | New treatment with renal replacement therapy in ICU | no-interaction treatment effect | Cox proportional hazards including the interaction between randomized group and septic shock, with adjustment for receipt of prehospital acid suppression |
|  | ICU mortality | no-interaction treatment effect | Cox proportional hazards including the interaction between randomized group and septic shock, adjusted for APACHE II score and receipt of prehospital acid suppression |
|  | Hospital mortality | no-interaction treatment effect | Cox proportional hazards including the interaction between randomized group and septic shock, adjusted for APACHE II score and receipt of prehospital acid suppression |
| **Analysis of Tertiary Outcomes (Septic shock vs. Others)** |  |  |  |
|  | Duration of ICU stay | no-interaction treatment effect | Linear regression including the interaction between randomized group and septic shock, adjusted for APACHE II score and receipt of prehospital acid suppression, with log-transformed ICU length of stay as the dependent variable. |
|  | Duration of hospital stay | no-interaction treatment effect | Linear regression including the interaction between randomized group and septic shock, adjusted for APACHE II score and receipt of prehospital acid suppression, with log-transformed hospital length of stay as the dependent variable. |
| **Sensitivity Analyses (Shock vs. Others)** |  |  |  |
| To determine if the effect of pantoprazole vs. placebo on the two primary outcomes differs in those patients with shock compared to those without shock | **Primary Efficacy Outcome:** Clinically important upper GI bleeding | no-interaction treatment effect | Cox proportional hazards including the interaction between randomized group and shock, with adjustment for receipt of pre-hospital acid suppression |
|  | **Primary Safety Outcome:** All-cause 90-day mortality | no-interaction treatment effect | Cox proportional hazards including the interaction between randomized group and shock, adjusted for APACHE II score and receipt of pre-hospital acid suppression |
| **Sensitivity Analyses (Septic shock of Pulmonary or Non-pulmonary origin vs. Others)** |  |  |  |
| To determine if the effect of pantoprazole vs. placebo on the 2 primary outcomes differs in those patients with septic shock due to pulmonary origin, compared to those patients with septic shock due to non-pulmonary | **Primary Efficacy Outcome:** Clinically important upper GI bleeding | no-interaction treatment effect | Cox proportional hazards including the interaction between randomized group and septic shock (pulmonary origin, non-pulmonary origin, no septic shock), with adjustment for receipt of pre-hospital acid suppression |
| origin, compared to those without septic shock | **Primary Safety Outcome:** All-cause 90-day mortality | no-interaction treatment effect | Cox proportional hazards including the interaction between randomized group and septic shock (pulmonary origin, non-pulmonary origin, no septic shock), adjusted for APACHE II score and receipt of pre-hospital acid suppression |

Abbreviations: APACHE, Acute Physiology and Chronic Health Evaluation; GI, gastrointestinal; ICU, intensive care unit.

Each analysis will include a subgroup interaction term to assess effect modification across groups. We considered each outcome and the hypothesis for each analysis is shown in **Table 1**. For example, a potential interaction could be that PPI-induced gastric pH elevation with antibiotic exposure in septic shock may promote dysbiosis and overgrowth of resistant pathogens that contribute to more secondary infections in PPI-exposed patients. However, this mechanism would depend on the combination of PPIs and antibiotics rather than PPIs alone, and remains speculative; therefore, we hypothesized that there would be no interaction regarding the secondary infectious outcomes. However, if an effect modification is identified for any outcomes, subgroup interaction will be evaluated using the ICEMAN instrument (26). All P-values will be two-sided, with P<0.05 considered statistically significant; aligned with our main report, for these subgroup analyses, we will use the sequential Holm–Šidak approach to adjust for multiple significance testing (27, 28). Analyses will be conducted using SAS software, version 9.4 (Cary, South Carolina).

We will conduct a sensitivity analysis examining patients with shock of any etiology, defined as receipt of vasopressors or inotropes at baseline regardless of admitting diagnosis, examining the primary efficacy and primary safety outcomes.

Although the cause of sepsis (pneumonia, urinary tract infection, or abdominal infection) was not associated with the rate of gastrointestinal bleeding in one 10-year U.S. database study (29), we will also conduct secondary subgroup analyses of patients with septic shock and pulmonary versus non-pulmonary sources to examine the effect of pantoprazole on the primary efficacy and primary safety outcomes.

### Ethics

The trial protocol was approved by ethics review boards at all participating sites. The consent models were either *a priori* informed consent or deferred consent (e.g., consent to continue); an opt-out model was used in one center.

## Discussion

This secondary analysis of the REVISE trial is a subgroup analysis that will examine consequences of SUP using intravenous pantoprazole versus placebo among patients with septic shock on the primary efficacy and safety outcomes of clinically important upper gastrointestinal bleeding and mortality within 90 days, as well as secondary trial outcomes.

Limitations of this study include its basis as a post hoc subgroup analysis potentially subject to type 1 error and spurious results. However, we will interpret findings conservatively. Patients will not be defined using SEPSIS-3 criteria (1) which were not available when this trial was launched; instead, we will employ an operational definition of septic shock using APACHE III admitting diagnostic categories reflecting infection with receipt of inotropes or vasopressors. Broad infectious etiologies were documented in the admitting diagnostic categories, COVID-19 status, and corticosteroid exposure, but no microbiological data or antibiotics were collected in the trial. This analysis may be underpowered to detect clinically important interaction effects, and the sample size will be smaller than pre-existing database and registry studies; however, the randomized blinded design reduces the risk of bias and confounding inherent in observational studies.

Strengths of this study will include minimization of bias due to the randomized exposure to pantoprazole or placebo, generating direct comparisons in the septic shock population. The clinically important upper gastrointestinal bleeding will be based on adjudication by two calibrated physicians who were blinded to treatment assignment (30). The extended subgroup analyses will offer randomized evidence on secondary infectious outcomes of concern for acid suppression among patients with septic shock. We have pre-specified no interaction treatment effect for any outcomes, after considering issues such as widespread antibiotic exposure (3), induced dysbiosis (31, 32), multifactorial bleeding diathesis (33), and the potential immunosuppressive effects of pantoprazole (34, 35). The generalizability of these findings will reflect the trial participation in 68 centers in 8 countries.

## Data Availability

This manuscript is a protocol and does not include data.

## Acknowledgement

We are grateful to Dr. Sylvain Lother for helpful suggestions on this manuscript.

